# Emulating a Target Trial to Evaluate the Impact of Empiric Antimicrobial Strategies on Mortality in Enterococcal Bloodstream Infections

**DOI:** 10.1101/2025.03.24.25324203

**Authors:** Juan Gago, Bo Shopsin, Victor J. Torres, Lorna E. Thorpe

## Abstract

Enterococcal bloodstream infections (BSI) caused by *Enterococcus faecalis* and *Enterococcus faecium* present significant public health concerns, particularly with rising vancomycin resistance (VRE). This study aimed to estimate the causal effect of two empiric strategies— immediate VRE coverage (daptomycin or linezolid) versus initial vancomycin or ampicillin with potential escalation—by emulating a target trial in electronic health records. We identified adult patients with positive blood cultures for *E. faecalis* or *E. faecium* from January 2017 to January 2023. Patients were assigned to immediate VRE coverage or initial VSE-active therapy with escalation within three days if clinical deterioration or new microbiologic data warranted. Using inverse probability weighting, we adjusted for baseline factors (e.g., age, comorbidities, prior antibiotic use) and time-varying confounders (changes in severity). Our intention-to-treat (ITT) and per-protocol analyses estimated 30-day all-cause mortality, with additional sensitivity analyses at 10 and 15 days. Among 799 patients, 45 (5.6%) received VRE-targeted therapy at baseline. The ITT analysis indicated a 1.35 percentage point mortality reduction (95% CI: –7.2% to 7.9%) with universal immediate VRE coverage, and the per-protocol analysis showed a risk difference of –3.76 points (95% CI: –14.9% to 20.4%). Sensitivity analyses yielded similar findings. In this population, immediate VRE coverage did not confer a clear survival advantage over standard therapy with potential escalation, supporting clinical equipoise and highlighting the need for larger trials (randomized or target trials) to evaluate optimal empiric regimens for enterococcal BSI.

## Introduction

Bloodstream infections (BSI) caused by *Enterococcus faecalis* and *Enterococcus faecium* are major clinical challenges due to their high rates of morbidity and mortality. Over the past three decades, these infections have become increasingly concerning due to the emergence of vancomycin-resistant *Enterococcus* (VRE) ^1,2^. Clinical management of VRE can be challenging since empiric antimicrobial treatment based on preliminary clinical assessments may not be active against the causative pathogen. Thus, empiric antimicrobial therapy protocols employed prior to pathogen identification and susceptibility test results may result increasingly inactive ^3,4^.

Different strategies have been proposed to improve treatments protocols of BSI, including shortening the time of empiric treatment by making a faster identification of the causative pathogen ^5–7^. Another potential intervention is broadening the spectrum of initial empiric antibiotics, and potentially including last-line antibiotics, such as linezolid or daptomycin. However, implementing these measures makes sense only if they are notably more effective in improving clinical outcomes compared to empiric treatment, and it is necessary to take into account the risks associated with their widespread adoption. Although they could improve the effectiveness of empiric treatment when AMR is unknown, widespread use of these last-line therapies may accelerate the development of antimicrobial resistance, potentially creating a bigger threat for patients and the community at large.

These complexities highlight the need for sound evidence backing the adoption of such measures. Conducting randomized control trials (RCTs) for these interventions presents considerable practical challenges, including logistical and financial constraints (requiring extensive resources for their design, execution, and follow-up phases). Enterococcal BSI is also a relatively rare event that mostly affects vulnerable patients with multiple co-morbidities. Many studies have pointed out the problem of generalizability of RCTs in these circumstances, where studies often have restrictive inclusion criteria that limit participant diversity ^8^. Altogether, these factors underscore the need to employ alternative study designs.

Observational studies have attempted to examine the impact different antimicrobial treatment strategies for enterococcal infections ^9,10^, but such non-randomized designs are often subject to design biases such as immortal time bias ^11–13^, which can result in misleading conclusions about the effectiveness of interventions ^14,15^. A limited number of studies have utilized a target trial emulation approach to examine antibiotic treatments for bacterial infections ^16–18^. This methodology structures observational data to mimic a hypothetical randomized experiment, facilitating a more actionable and interpretable research question, especially in clinical settings. It also prevents immortal time, by establishing a clear ‘time zero’. Estimation of the observational analog of the intention-to-treat effect requires adjustment for time-fixed (baseline) confounders and estimation of the per-protocol effect requires further adjustment for time-varying confounders, which can be appropriately achieved via g-methods ^19^ even in the presence of treatment-confounder feedback (e.g., if antimicrobial treatment affects clinical severity and severity informs the election of subsequent antimicrobial treatment).

Improving empiric treatment for BSI, such it could be incorporating antimicrobials with anti-VRE activity, is particularly important as these infections present several unique clinical challenges. These include high associated mortality and a higher propensity for occurrence among patients from vulnerable populations. Furthermore, enterococcal resistance patterns vary significantly across different locations and populations ^20^ and effectiveness of antimicrobial treatment protocols are linked to the distribution of pathogens’ AMR. Thus, it is crucial to address the impact of changes in the protocols of antimicrobial treatment on enterococcal BSI utilizing granular local data.

This study aims to evaluate two different antimicrobial treatment strategies: one including antibiotics that cover VRE (linezolid or daptomycin) as treatment from day 0. The other strategy is the more common scenario involving use of agents that cover enterococcus but not VRE (ampicillin or vancomycin), and escalating to daptomycin or linezolid when data on susceptibility is available or if clinical conditions worsen, among patients with a diagnosis of enterococcal infection. Using electronic health records (EHR) from a large New York City healthcare system, our study employs a causal inference-informed target trial emulation model, as outlined by Hernan and Robins ^19^, to evaluate the impact of these antibiotic treatment strategies on all-cause mortality at 30 days as main outcome. This approach contributes to a growing body of research applying causal inference methods to observational data on acute infectious diseases and AMR. It provides valuable insights into a clinically relevant question regarding the management of enterococcal infections, especially in the context of changing VRE epidemiology.

## Methods

We aim to answer the research question by specifying the protocol of a (hypothetical) pragmatic randomized trial, or the target trial, then emulating this target trial using observational data.

### Specification of the Target Trial

**In our target trial, we want** to compare the effect of employing agents that cover VRE (linezolid or vancomycin) from day 0 against administration of agents that cover vancomycin-sensitive Enterococcus (VSE) (i.e. vancomycin or ampicillin) and subsequent adjustment based on deterioration of clinical condition or new information from culture or susceptibility test. The components of the protocol are summarized in **Table 1**.

**Table 1.**
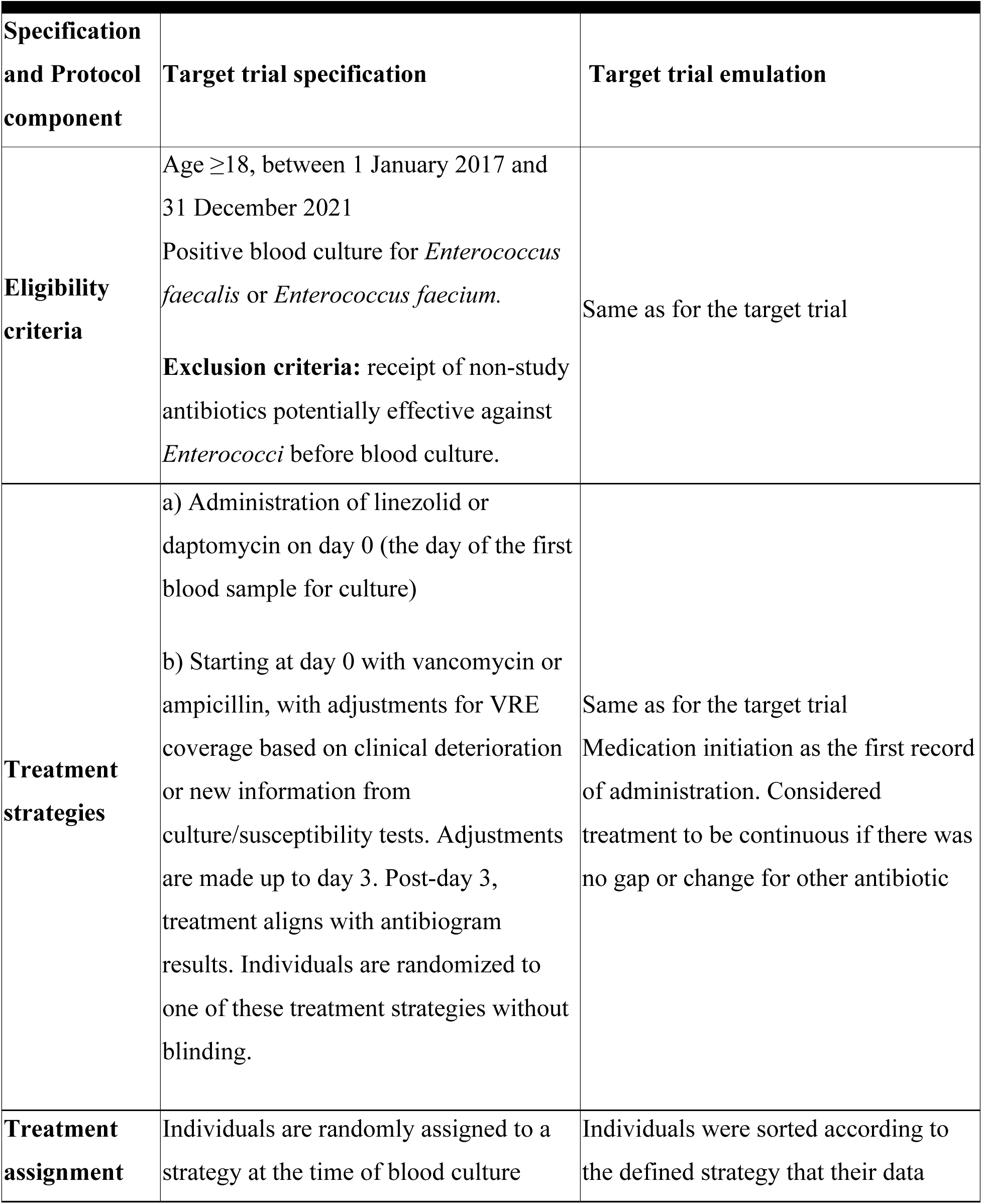
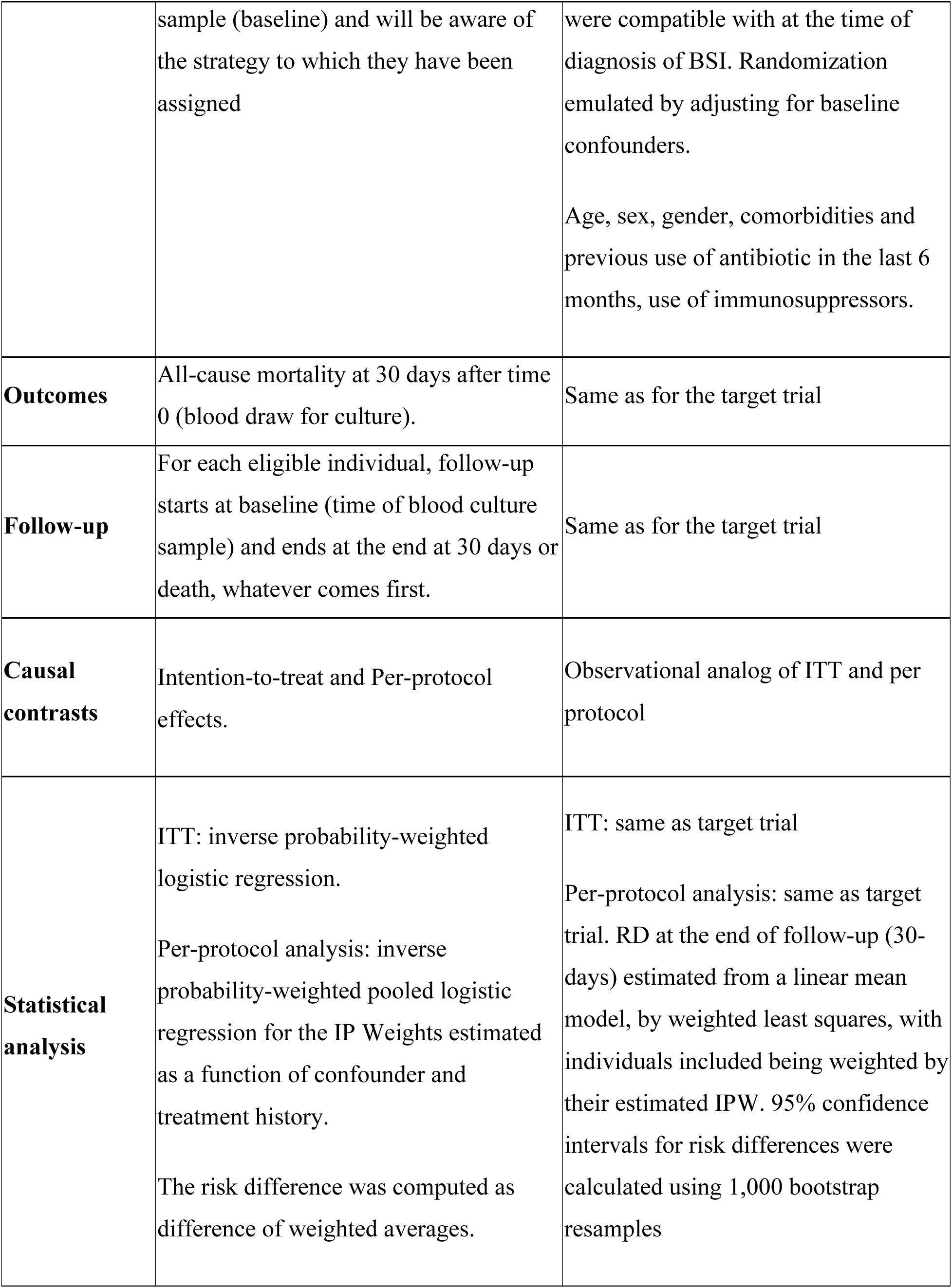
Specification of Target Trial and emulation.

| **Specification and Protocol component** | **Target trial specification** | **Target trial emulation** |
| --- | --- | --- |
| **Eligibility criteria** | Age $\geq 18$, between 1 January 2017 and 31 December 2021<br>Positive blood culture for *Enterococcus faecalis* or *Enterococcus faecium*.<br>**Exclusion criteria:** receipt of non-study antibiotics potentially effective against *Enterococci* before blood culture. | Same as for the target trial |
| **Treatment strategies** | a) Administration of linezolid or daptomycin on day 0 (the day of the first blood sample for culture)<br>b) Starting at day 0 with vancomycin or ampicillin, with adjustments for VRE coverage based on clinical deterioration or new information from culture/susceptibility tests. Adjustments are made up to day 3. Post-day 3, treatment aligns with antibiogram results. Individuals are randomized to one of these treatment strategies without blinding. | Same as for the target trial<br>Medication initiation as the first record of administration. Considered treatment to be continuous if there was no gap or change for other antibiotic |
| **Treatment assignment** | Individuals are randomly assigned to a strategy at the time of blood culture | Individuals were sorted according to the defined strategy that their data |
|  | sample (baseline) and will be aware of the strategy to which they have been assigned | were compatible with at the time of diagnosis of BSI. Randomization emulated by adjusting for baseline confounders.<br>Age, sex, gender, comorbidities and previous use of antibiotic in the last 6 months, use of immunosuppressors. |
| **Outcomes** | All-cause mortality at 30 days after time 0 (blood draw for culture). | Same as for the target trial |
| **Follow-up** | For each eligible individual, follow-up starts at baseline (time of blood culture sample) and ends at the end at 30 days or death, whatever comes first. | Same as for the target trial |
| **Causal contrasts** | Intention-to-treat and Per-protocol effects. | Observational analog of ITT and per protocol |
| **Statistical analysis** | ITT: inverse probability-weighted logistic regression.<br>Per-protocol analysis: inverse probability-weighted pooled logistic regression for the IP Weights estimated as a function of confounder and treatment history.<br>The risk difference was computed as difference of weighted averages. | ITT: same as target trial<br>Per-protocol analysis: same as target trial. RD at the end of follow-up (30-days) estimated from a linear mean model, by weighted least squares, with individuals included being weighted by their estimated IPW. 95% confidence intervals for risk differences were calculated using 1,000 bootstrap resamples |

Eligibility criteria would include patients aged 18 and older with a laboratory confirmed diagnosis of *E. faecalis* or *E. faecium* bacteremia for the period January 2017 to January 2023. Patients should not have a concurrent active infection or be receiving antibiotics potentially effective against the enterococcus. Patients would be excluded if they had records of using antibiotics in the previous 7 days to admission. We would include patients with independent events of enterococcal BSI (i.e., not occurring during the same admission, and not recorded as the result of previous treatment failure).

The interventions of interest would be: a) the administration of linezolid or daptomycin on day 0 (the day of the first blood sample for culture), compared to b) administration of vancomycin or ampicillin on day 0, with adjustments for VRE coverage based on clinical deterioration or new information from culture/susceptibility tests. Adjustments would be made up to day 3. Post-day 3, treatment would align with antibiogram results. Individuals would be randomized to one of these treatment strategies without blinding.

The outcome of interest is 30-day all-cause mortality. For each eligible participant, follow-up starts when the first blood sample for blood culture is obtained and ends after 30 days or when the patient died, whichever occurs first. We would perform a sensitivity analysis of the effect of the treatment on early mortality (10-day and 15-day all-cause mortality). This sensitivity analysis with different risk periods would allow to evaluate if there are early effects of the treatment on mortality The intention-to-treat effect (ITT) and the per-protocol effect are our causal contrasts of interest. The ITT is starting antibiotics with VRE coverage (linezolid or daptomycin) vs. starting antibiotics covering VSE pathogens (vancomycin or ampicillin). The per protocol effect is the effect of continuous VRE-covering antibiotic treatment vs. the effect of initiating with VSE-covering antibiotics and adjusting accordingly based on clinical severity changes or information from blood culture.

For the per-protocol analysis, we would assume that revision of empiric treatment followed a current standard-of-care approach, where empiric treatment is revised under two scenarios: 1) when blood culture results provide new information regarding the causal pathogen (e.g. pathogen species and antimicrobial susceptibility results) or 2) when a patient’s clinical parameters deteriorate. Switching (escalating) antimicrobials before receiving blood culture results is generally not advised by stewardship guidelines unless clinicians consider it necessary due to clinical deterioration. There are no exact indicators for when to escalate antimicrobials before pathogen identification. ICU admission serves as a reliable indicator of clinical deterioration, as patients admitted to intensive care have undergone thorough evaluation. Furthermore, the decision for ICU admission often reflects an expert opinion that the patient requires closer monitoring. This event is always documented in the EHR. Taking this approach into consideration, ICU admission post-blood culture would be used as time-varying indicator of clinical deterioration. **Fig. 1** is a direct acyclic graph (DAG) representing the different variables and causal relationships considered for this study.

**Fig. 1.**
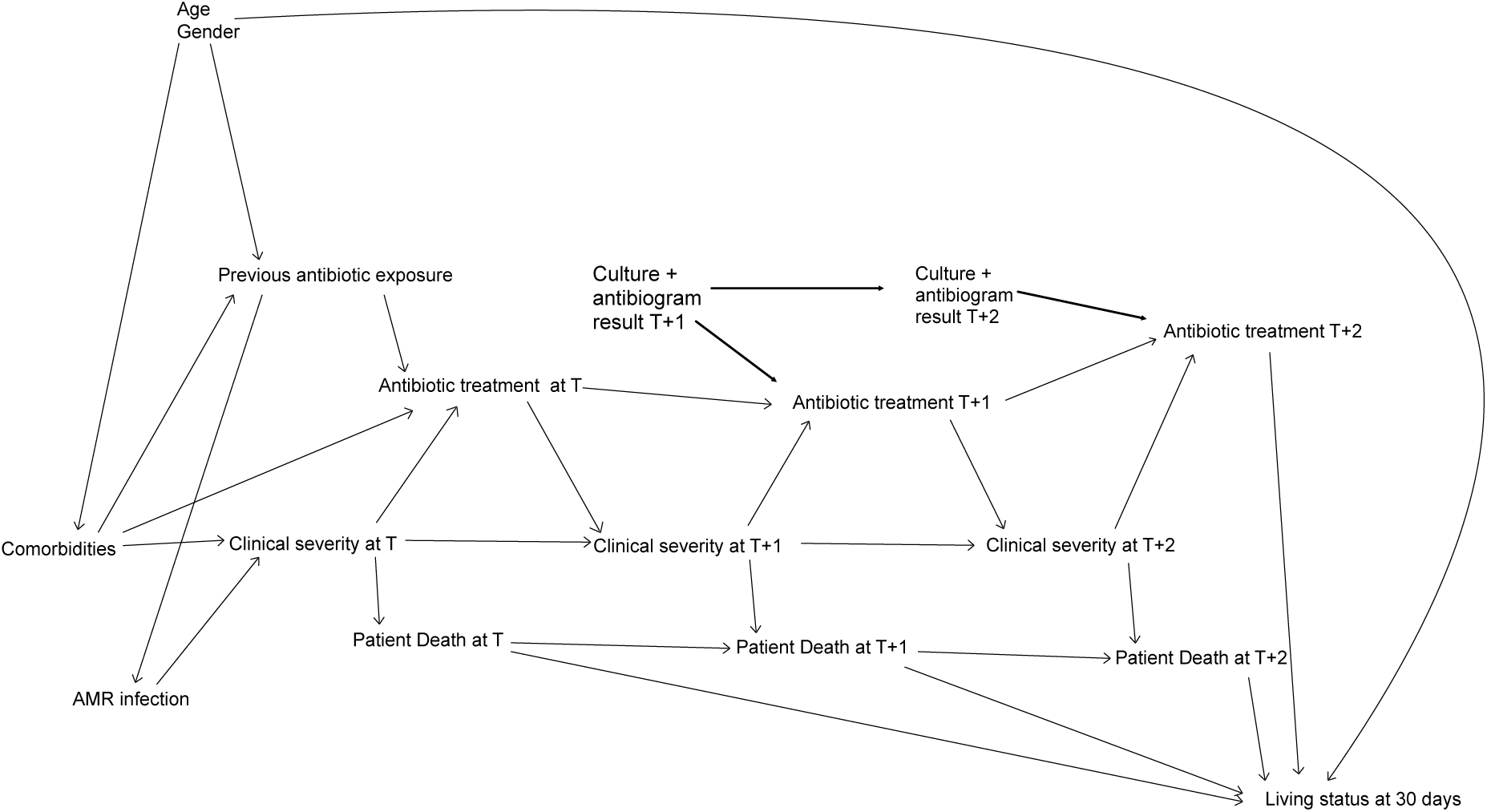
Direct Acyclic Graph (DAG) illustrating variables implied in the treatment of enterococcal BSI and the evaluated outcome of mortality at 30 days.

### Emulation of the target trial

We emulated this target trial using the EHR system at NYU Langone Health, with data from inpatient services located in two hospitals in the boroughs of Manhattan and Brooklyn. Patients’ diagnosis of enterococcal BSI was identified with the result of a positive blood culture for either *E. faecalis* or *E. faecium*. Antibiotic treatments were identified as documented administration of any of the drugs of interests. We summarized the timeline of events for patients with BSI in **Fig. 2**. The outcome of interest was the same that in the target trial, all-cause 30-day mortality, defined as a death within 30 days after the first blood sample was extracted for blood culture. As sensitivity analyses, we used as outcomes 10-day and 15-day mortality to capture potential early effects of treatment. This sensitivity analysis with different risk periods also is an aide to assess the potential residual time-varying confounding (i.e., early complications that might affect the choice of antimicrobial treatment and the outcome). In-hospital mortality was directly recorded in the system. Deaths outside the hospital setting are reported by each State to the Social Security Administration. NYU Langone’s records are routinely cross-referenced with these reports, which ensures minimal misclassification of the outcome. Given that all participants were inpatients, there was negligible loss-to-follow-up in our study. All patients for whom a blood sample was drawn for blood culture are eventually diagnosed, which allowed us to make the assumption of instant diagnosis at time 0. Treatment administration and any subsequent events were automatically documented in the EHR.

**Fig. 2.**
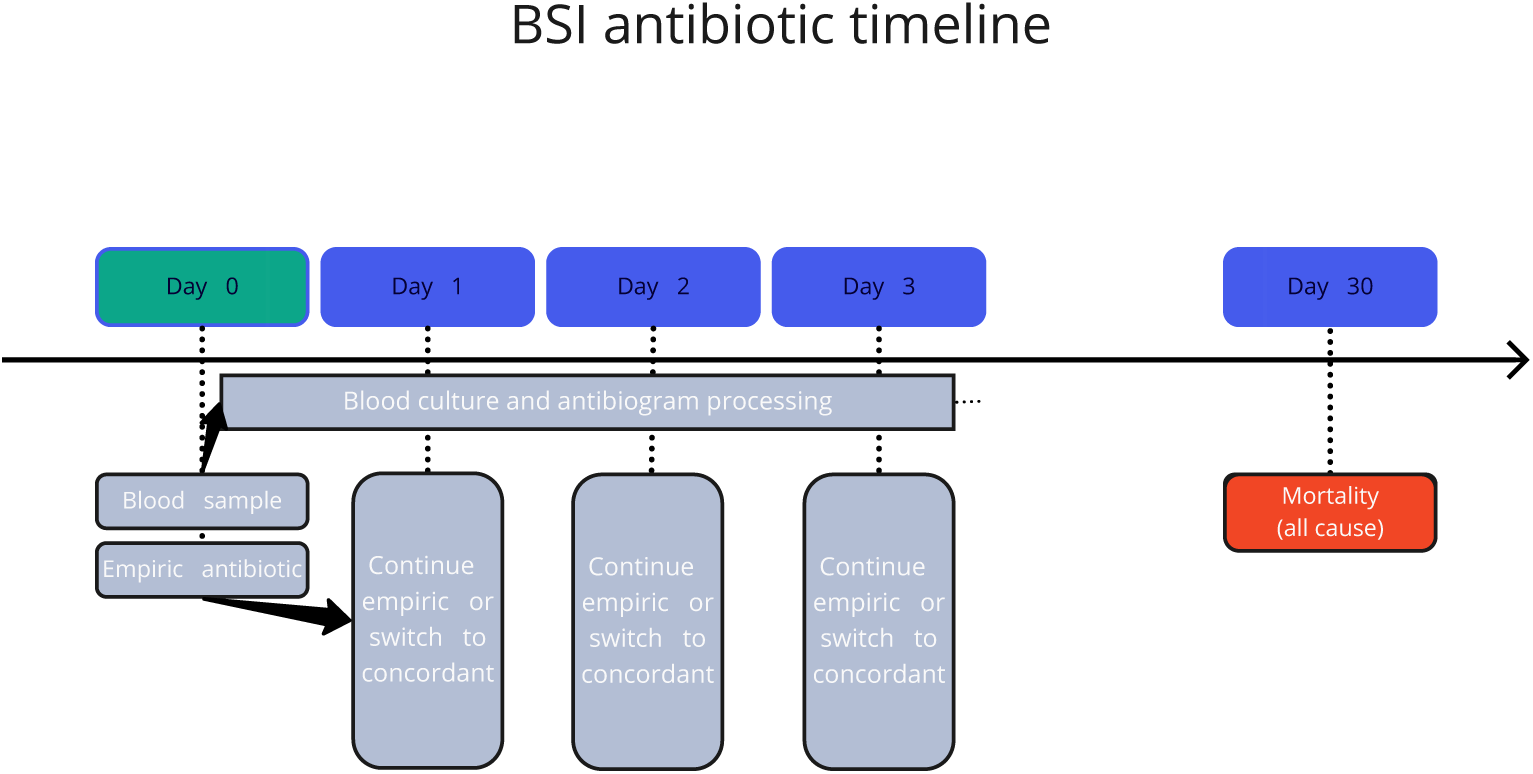
Timeline of events for patients with BSI. Simplified timeline of events following an admission with a diagnosis of bloodstream infection.

The observational analog of the intention-to-treat (ITT) effect was estimated as follows: We first used a logistic regression model to estimate the probability of being assigned to each treatment at baseline. This regression included the following variables: (BSI), including age (accounted for with both linear and quadratic terms), sex, race, ethnicity, history of antibiotic use (documented as the use of antibiotics up to one week prior to the BSI event), previous BSI events, the Charlson comorbidity index, ICU admission status at baseline, and history of immunosuppression. These probabilities were then utilized to estimate IP weights. Finally, we applied the IP weights in a weighted linear model, which estimated the risk differences (RD). To mitigate the influence of outliers, these weights were truncated at the 99th percentile. For the estimation of the per-protocol effect we performed the following steps: we calculated IP weights based on the probability of patients adhering to their assigned treatment at each time point when treatment adjustments were allowed (from day 1 to day 3). These weights were estimated using baseline confounders as described earlier, along with time-varying confounding factors including ICU admission as a proxy of severity, and the patients’ treatment history. A description of the variables included to calculate the weights and how these were coded in the models are shown in in **Table 2**.

**Table 2.** Variables used for the construction of weights.

| Variable name | Description | Format | Coding |
| --- | --- | --- | --- |
| **Time invariant variables** |  |  |  |
| *Age* | Age at baseline in years | numeric |  |
| *Gender* | Gender | categorical | male<br>female |
| *History of antibiotic use* | Record of antibiotic use in the previous year up to a week before diagnosis | categorical | no / yes |
| *Charlson comorbidity index* | Measure of comorbidities at baseline | numeric |  |
| *Previous episodes of BSI* | number of previous events of BSI considered independent from each other | numeric |  |
| *Immunosuppressed status* | Record of immunosuppression due to medication or base condition | categorical | no / yes |
| **Time variant variables** |  |  |  |
| *Admission to ICU* | Admission to ICU as proxy for deterioration of clinical status | categorical | no / yes |

Risk differences were calculated fitting a linear mean model, by weighted least squares, with individuals included being weighted by their estimated IP weights as described above. 95% confidence intervals for risk differences were calculated using 1,000 bootstrap resamples. All analyses were conducted using R software version 4.1.0.

## Results

A total of 799 patients were included in our study, and they were categorized based on antibiotic treatment initiated on day 0. We present baseline characteristics in **Table 3**. Of these patients, only 45 (5.6%) received daptomycin or linezolid as their first option on day 0, while the rest started with either vancomycin or ampicillin. Of these, 66 (8.2%) switched to VRE-covering antibiotics on day 1, 89 (11.1%) on day 2, 69 (8.6%) on day 3, and 458 (57.3%) remained on their initially-assigned antibiotic. Nearly 1 in ten patients (9.0%) switched to a different antibiotic treatment. The cohort was predominantly male (63.1%) and white (60.2%), with variations in age and race observed across the different delay groups. The mean Charlson comorbidity index was slightly higher in the group assigned to daptomycin or linezolid (6.6), indicating a potentially higher burden of comorbidities.

**Table 3.** Characteristics of the patients classified by antibiotic treatment at time 0.

|  | Initiating VSE<br>antimicrobials<br>(N=754) | Initiating VRE<br>antimicrobials<br>(N=45) | Total (N=799) |
| --- | --- | --- | --- |
| **History of Antibiotic Treatment** |  |  |  |
| Yes | 459 (60.9%) | 40 (88.9%) | 499 (62.5%) |
| **Pathogen Species** |  |  |  |
| *Enterococcus faecalis* | 510 (67.6%) | 22 (48.9%) | 532 (66.6%) |
| *Enterococcus faecium* | 244 (32.4%) | 23 (51.1%) | 267 (33.4%) |
| **Sex** |  |  |  |
| Female | 278 (36.9%) | 19 (42.2%) | 297 (37.2%) |
| Male | 476 (63.1%) | 26 (57.8%) | 502 (62.8%) |
| **Race** |  |  |  |
| Black | 81 (10.7%) | 6 (13.3%) | 87 (10.9%) |
| Asian | 70 (9.3%) | 3 (6.7%) | 73 (9.1%) |
| White | 454 (60.2%) | 28 (62.2%) | 482 (60.3%) |
| Other<sup>1</sup> | 149 (19.8%) | 8 (17.8%) | 157 (19.6%) |
| **Ethnicity** |  |  |  |
| Hispanic | 78 (10.3%) | 5 (11.1%) | 83 (10.4%) |
| Not of Spanish/Hispanic Origin | 605 (80.2%) | 38 (84.4%) | 643 (80.5%) |
| Prefer not to answer | 71 (9.4%) | 2 (4.4%) | 73 (9.1%) |
| **Age (Years)** |  |  |  |
| Mean (SD) | 70.2 (16.2) | 59.1 (20.9) | 65.5 (20.6) |
| **Number of previous BSI events** |  |  |  |
| Mean (SD) | 1.0 (1.7) | 1.2 (2.5) | 1.0 (1.7) |
| **Immunosuppression history** |  |  |  |
| Yes | 104 (13.8%) | 10 (22.2%) | 114 (14.3%) |
| **Charlson Score Index** |  |  |  |
| Mean (SD) | 4.3 (3.7) | 6.6 (4.8) | 4.5 (3.8) |

*Note.*<sup>1</sup> Categories with less than 2 patients were collapsed into others, including Native Americans and Native Pacific Islanders.

Regarding pathogen distribution, *E. faecalis* was the most common (66.6%) overall. *E. faecalis* and *E. faecium* were similarly distributed among those assigned VRE-covering antibiotics from day 0 (E. faecalis 48.9%, E. faecium 51.1%).

In the emulation of our target trial, comparing the strategies of starting with daptomycin or linezolid from time 0 – when the first blood sample for culture was drawn – to starting with vancomycin or ampicillin, we found that had all the participants had started with VRE coverage treatments, the 30-day mortality observed would have decreased, being the RD –1.35% (95% CI: –7.2% to 7.9%) (**Table 4**).

**Table 4.** Estimates of Risk Differences comparing the strategies initiating daptomycin or linezolid vs. initiating vancomycin or ampicillin.

|  | Risk difference | 95% CI |  |
| --- | --- | --- | --- |
|  |  | Lower bound | Upper bound |
| Intercept | 21.3% | 18.3% | 22.9% |
| *Antimicrobial treatment initiated* |  |  |  |
| Vancomycin or Ampicillin | Ref. | - | - |
| Daptomycin or Linezolid | - 1.35% | -13.0% | 22.9% |

*Note.* RD: Risk difference.

When comparing the results of using daptomycin or linezolid from time 0 versus starting with vancomycin or ampicillin and then switching to VRE-covering antibiotics on different days (**Table 5**), the risk difference between sustained VRE-covering antibiotics and the dynamic strategy was –3.76% (95% CI: –14.9% to 20.4%). These data are compatible with a clinical equipoise between VRE-covering antibiotics from day 0 and the standard of care for the treatment of enterococcal BSI.

**Table 5.** Estimates of Risk Differences comparing the strategies of starting and continuing with daptomycin or linezolid vs. starting vancomycin or ampicillin and adjusting treatment as indicated.

|  | Risk difference | 95% CI |  |
| --- | --- | --- | --- |
|  |  | Lower bound | Upper bound |
| Intercept | 18.8% | 7.49% | 27.6% |
| *Antimicrobial treatment* |  |  |  |
| Daptomycin or Linezolid from day 0 | Ref. | - | - |
| Starting Vancomycin or ampicillin, switching to VRE-covering | 3.76% | -14.9% | 20.4% |

*Note.* RD: Risk difference.

In the sensitivity analysis to estimate mortality occurring earlier in the course of the episode of BSI, we observed the following: 7 out of 45 patients in the VRE-covering group died within the first 10 days. From day 10 to day 15 no new deaths occurred in this group. The risk difference for mortality at 10 days after baseline comparing the two treatment strategies was 4.2% (95% CI: – 5.2% to 16.9%) had all the patients received VRE-covering antibiotics. For mortality at 15 days, the estimated risk difference was 1.7% (95% CI: –7.9% to 14.7%) had all the patients received VRE-covering antibiotics.

## Discussion

Our study explored the effect of different antibiotic treatment strategies on 30-day all-cause mortality in patients with BSI caused by Enterococcus. We found that the estimates of the effect of the early introduction of antimicrobial agents covering VRE on 30-day mortality risk in patients diagnosed with enterococcal BSI are compatible with clinical equipoise compared to the standard of care. Despite the wide confidence intervals in our estimates, indicating the imprecision of the point estimates, this study represents an initial attempt to answer our causal question: whether introducing new agents when antimicrobial resistance is unknown in enterococcal infections could improve clinical outcomes among these patients. Further studies on larger datasets could provide more precise answers regarding the average treatment effect of this strategy. The imprecise estimates can, in part, be explained by the fact that initiating treatment with VRE-covering agents is not a common event. When estimating earlier mortality, the risk difference for mortality at day 10 and day 15, we also observed data compatible with clinical equipoise had all the patients in our study been treated with VRE-covering agents.

Protocols in managing enterococcal BSI might need to be evaluated based on local AMR distribution; thus, we introduce a piece of evidence that can help evaluate antimicrobial treatment strategies in clinical settings. A careful evaluation of specific interventions, such as the early use of daptomycin or linezolid for patients at high risk of resistant strains, might be important steps in order to inform changes in clinical practice. As these data are compatible with clinical equipoise between the strategies, they represent an important piece of evidence in favor of conducting further evaluation of these treatments in future clinical trials.

The rise in VRE prevalence has become a significant challenge in the last decades ^21^, necessitating new considerations when assessing therapeutic options. Previous studies have focused on the timing of antibiotic treatment of enterococcal BSI, such as the work of Zasowski et al. (2016), who used a non-parametric regression tree model to evaluate this question, and Russo (2022) who centered on the association of risk factors to 30-day mortality. Earlier studies ^14,15^ have found contradictory answers to the problem of timing of concordant antibiotic administration, most likely due to non-optimal adjustment for confounding or the presence of immortal time bias. The use of causal models such as target trial emulations to evaluate antibiotic treatment has not yet been widely implemented, although there are precedents in the important work of Lim et al. (2021) evaluating the delay of antimicrobial use treating Acinetobacter, as well as Evans et al. (2022) assessing the effect of duration of antibiotic treatment for *S. aureus,* and Pouwels et al. (2017) evaluating appropriate antibiotic treatment in patients with Enterobacteriaceae BSI in the ICU. The heterogeneity of AMR distribution, as well as the rapidly changing landscape of pathogens, underscores the importance of having reliable evidence available to improve treatment strategies, stewardship guidelines, and other measures of infection control. Particularly for enterococcal BSI, we have limited therapeutic options available for VRE (linezolid and daptomycin being the remaining last-line antibiotics employed). In clinical challenges like this, characterized by high mortality and morbidity, it is crucial to accurately assess the effectiveness of medical interventions and treatments that can improve patients’ outcomes.

The use of molecular PCR tests for pathogen identification has emerged as an important tool to reduce the time to get treatment for AMR strains. One such test is the BCID2 Panel, a molecular diagnostic tool that identifies a wide array of pathogens and antibiotic resistance genes associated with bloodstream infections, and it can significantly expedite the switch from empirical treatment to concordant antibiotics, when necessary (Banerjee et al., 2015). These tests are performed once a blood culture turns positive and can reduce the pathogen identification time to 24-48 hours. Nevertheless, challenges persist. While PCR tests offer faster results, they are not yet universally adopted. Health systems with limited resources find adoption challenging and costly. In places where the PCR test is employed for detecting pathogen species and resistance genes, inherent delays can still exist ^22^. Current technology only permits tests on positive cultures, not directly from blood samples (as it is done for viral detection techniques). This means there is often a one or two-day gap between blood sample collection and results, delaying the potential switch in antimicrobials. Adjust empiric treatments and incorporating a last-line agent with broader coverage for enterococcal infections such as daptomycin or linezolid in places with a high prevalence of VRE has not been widely evaluated in clinical settings ^23^. However, decisions come with considerations. For instance, changing empirical regimes to cover VRE might inadvertently increase the misuse of this antibiotic. This can, in turn, escalate the circulation of resistant strains, exacerbating the issue. In scenarios where VRE is more prevalent, multiple strategies are needed to improve the antibiotic treatment strategies.

Our study has a few important limitations. First, data from electronic health records are routinely collected for clinical purposes and are not designed for research purposes. Records of previous antibiotic use are imperfect, most likely not including antibiotics taken as outpatients or if the person is in a long-term care facility. In this study, we assumed that data on the timing of treatments and outcomes were accurately recorded and that the dataset extracted was reliable. We also assumed that measured covariates were sufficient to adjust for confounding factors (**Table 2**). In the present study of inpatients, we also assumed that selection bias due to loss to follow-up was non-existent. We also assumed that the models were correctly specified, however, the weighting process and parametric models employed in our study may have been subject to misspecification. Additionally, the use of IPW can result in instability due to high variance stemming from extreme weights. Further studies would be needed to assess different antibiotic strategies for the treatment of enterococcal BSI, being the gold standard, for this scenario, an RCT assessing the effect of changing antibiotic regimes in clinical outcomes. However, as previously noted, resource constraints make conducting this type of study challenging. Lastly, our estimates include wide confidence intervals, and thus further studies with greater precision are needed. However, as pointed out by Hernán <u>(2022)</u>, pursuing studies, even under the possibility of imprecise outcomes, is vital when tackling significant causal questions. Avoiding observational studies due to concerns about being ‘underpowered’ hinders the growth of scientific evidence. Decisions about conducting such studies should be based on factors such as the causal question’s importance, resource allocation priorities, or the potential of new insights on the problem, rather than on expected precision. If feasible, pooling results with similar studies trying to answer the same causal question can pave the way to obtaining more precise answers. Furthermore, observational studies like this can also act as preliminary evidence until randomized trials provide more definitive answers.

Our study provides some evidence by organizing data in a target trial emulation and employing g-methods to control for time-varying confounding. This framework allows a thorough analysis of observational data and effectively addresses potential biases introduced in the design of other observational studies. Focusing on the critical healthcare issue of enterococcal BSI, our work offers valuable insights on the use of clinical data combined with a causal framework in order to asses hospital antimicrobial policies and strategies.

## Conclusion

Our study highlighted the impacts of different antibiotic treatment strategies on mortality among patients with enterococcal BSI. With current increasing trends of VRE BSI and well documented challenges in its treatment, there is a pressing need for tools evaluating potential changes in empiric antimicrobial therapies. The use of EHR in clinical settings with the appropriate methods can be powerful in guiding interventions to strengthen antimicrobial stewardship and improve outcomes when dealing with AMR.

## Data Availability

All data produced in the present study are available upon reasonable request to the authors after proper de-identification.

